# Predicting COVID-19 Infection Among Older Syrian Refugees in Lebanon to inform outbreak preparedness: A Multi-Wave Survey

**DOI:** 10.1101/2024.01.17.24301436

**Authors:** Berthe Abi Zeid, Tanya El Khoury, Sawsan Abdulrahim, Hala Ghattas, Stephen McCall

## Abstract

**Background:** Older refugees, exposed to a cluster of biological and social vulnerabilities, are more susceptible to infectious diseases outbreaks and their complications. This study contributes to health emergency preparedness efforts by developing and internally validating a predictive model estimating COVID-19 infection risk among older Syrian refugees in Lebanon using social determinants. Additionally, it described the barriers to PCR testing among those who reported a COVID-19 infection.

**Methods:** This was a cross-sectional analysis of a five-wave longitudinal study conducted between September 2020 and March 2022. Syrian refugees aged 50 years or older living in households that received assistance from a humanitarian organization were interviewed by phone. Self-reported COVID-19 infection was the outcome of interest. The predictors were identified using adaptive LASSO regression. The model performance and discrimination were presented using the calibration slope and the Area Under the Curve.

**Results:** Of 2,886 participants (median [IQR] age: 56[52-62]; 52.9% males), 283 individuals (9.8%) reported a COVID-19 infection at least once. Six predictors for COVID-19 infection were identified: living outside informal tented settlements, having elementary and preparatory education or above, having chronic conditions, not receiving cash assistance, being water insecure and having unmet waste management needs. The model had moderate discrimination and good calibration. Nearly half of the cases were diagnosed through PCR testing. The main reasons for not testing were perception that the tests were unnecessary (n=91[63.6%]) or inability to afford them (n=46[32.2%]).

**Conclusions:** High-risk individuals should be targeted based on predictive models incorporating social determinants. Implementing awareness campaigns, screening measures, and cash assistance may reduce vulnerability in future pandemics.

## 1. Introduction

The COVID-19 pandemic was one of the most recent global emergencies in modern history and its long-term impact continues to shape health systems and economies worldwide.^1^ As of June 15, 2025, the World Health Organization has recorded a total of 778,240,931 confirmed cases of COVID-19, encompassing 7,097,572 deaths, globally.^2^ According to Oxfam’s report published in March 2022, the COVID-19 mortality rate has been four times higher in lower-income countries than those with higher incomes.^3^ In addition, wealth inequalities in the incidence of COVID-19 infections have been reported globally.^4^ COVID-19 pandemic exacerbated preexisting health inequalities, further marginalizing vulnerable communities by restricting accessibility to essential services.^5^ As the world transitions to recovery, COVID-19 can be a critical case study in pandemic preparedness and anticipatory action, highlighting the urgent need to strengthen global and local capacities for mitigating future pandemic threats, particularly among marginalized populations.

Displaced populations have increased exposure to infectious diseases and are disproportionally impacted by ‘shocks’ such as the COVID-19 pandemic.^6^ For instance, refugees face various difficulties such as overcrowding, shortages of supply of hygiene products and personal protective equipment, limited access to healthcare services, fear of deportation, and cultural and language barriers.^7^ Moreover, older refugees have a higher prevalence of pre-existing chronic illnesses compared to both the host community and younger adults, putting them at higher risk of severe morbidity and mortality during health emergencies.^8^

During the COVID-19 pandemic, humanitarian organizations worldwide encountered substantial obstacles due to containment measures and nationwide lockdowns, which limited their ability to aid and protect vulnerable populations.^9^ The UN High Commissioner for Refugees (UNHCR) declared a funding shortfall of USD 255 million in its plan for COVID-19 prevention and response among refugee populations.^10^ Consequently, humanitarian assistance was limited in many settings, which may have exposed refugee populations disproportionally to infection risk through reduced ability to self-isolate due to the need to receive a livelihood.^7^

In 2024, 73% of the 123.2 million forcibly displaced people lived in low-and middle-income countries.^11^ Lebanon hosts the highest number of refugees per capita, including 1.5 million Syrian refugees, of whom 831,053 are officially registered with the UNHCR.^12^ In addition to the multiple economic, political and health crises in Lebanon, thousands of Syrians have been forced to return to Syria, creating an environment of fear among Syrians living in the country, regardless of their refugee status.^13^ These combined challenges have deteriorated the living conditions of Syrian refugees and are associated with loss of livelihoods, thereby placing them at an elevated risk of infection and its impacts.^7^ Despite their high susceptibility to COVID-19 infection, testing in this population has remained limited with a major barrier being lack of affordability.^14^

Understanding the social determinants of infectious disease amongst vulnerable populations is crucial for designing effective preparedness and response strategies, particularly in the early stages of future pandemic, when information is limited and rapid decisions are needed. Prediction models can help humanitarian and public health actors to efficiently identify individuals most as risk of infection, enabling them to provide essential assistance for prevention, while allocating limited resources more equitably.

Previous studies conducted among patients who were tested for COVID-19 in the United States have identified age, gender, educational attainment, marital status, housing conditions, socioeconomic status, and presence of chronic illnesses as predictors of COVID-19, alongside a broader set of biological and clinical factors (e.g., biomarkers).^15,16^ These factors might have an impact on an individual’s capacity to seek healthcare, comply with treatment, and observe physical distancing measures.^15^ However, without accounting for the combined contribution of the remaining biological and clinical predictors in the models, these social determinants alone may not be sufficient to accurately identify individuals at highest risk, particularly in the early stages of a pandemic, when clinical and diagnostic data are limited. This underscores the need to evaluate the predictive value of social determinants on their own through dedicated models. In addition, to date, there has been a scarcity of predictive models to assess the risk of infection among refugees or older adults in the Middle East and North Africa (MENA) region. Therefore, this study aimed to develop and internally validate a predictive model to estimate the risk of COVID-19 infection among older Syrian refugees in Lebanon. It also aimed to describe the barriers to PCR testing among those who had COVID-19 infection. These findings contribute to health emergency preparedness efforts by providing an actionable model and approach that can be used to identify populations at risk in future pandemics and can be adapted to other settings or infectious diseases.

## 2. Methods

### 2.1. Study design and setting

This was a cross-sectional analysis of a five-wave longitudinal study that monitored the vulnerabilities of older Syrian Refugees residing in Lebanon to COVID-19 over time. The study complied with the Transparent Reporting of a Multivariable Prediction Model for Individual Prognosis or Diagnosis (TRIPOD) reporting guideline and Strengthening the Reporting of Observational Studies in Epidemiology (STROBE) reporting guideline for prediction modelling.^17,18^

### 2.2. Sampling and study population

This study targeted Syrian refugees aged 50 years or older who were identified from a complete roster of beneficiary households of a humanitarian Non-Governmental Organization (NGO) [Norwegian Refugee Council]. Within the beneficiary sample listing, all households in Lebanon that received assistance from the humanitarian organization between 2017 and 2020 and included an age-eligible member, were contacted by phone (n=17,384). Adults aged 50 years or older were invited to participate in the study. In households where multiple eligible adults were present, one individual was randomly selected. Verbal informed consent was taken from all participants and an evaluation of their capacity to consent was conducted for those aged 65 years or older.^19^ The same participants were contacted over the course of five waves, spanning from September 22, 2020, to March 14, 2022. The study population for the current analysis, as depicted in Figure 1, consisted of participants who completed the first, second, and fifth waves.

**Figure 1.**
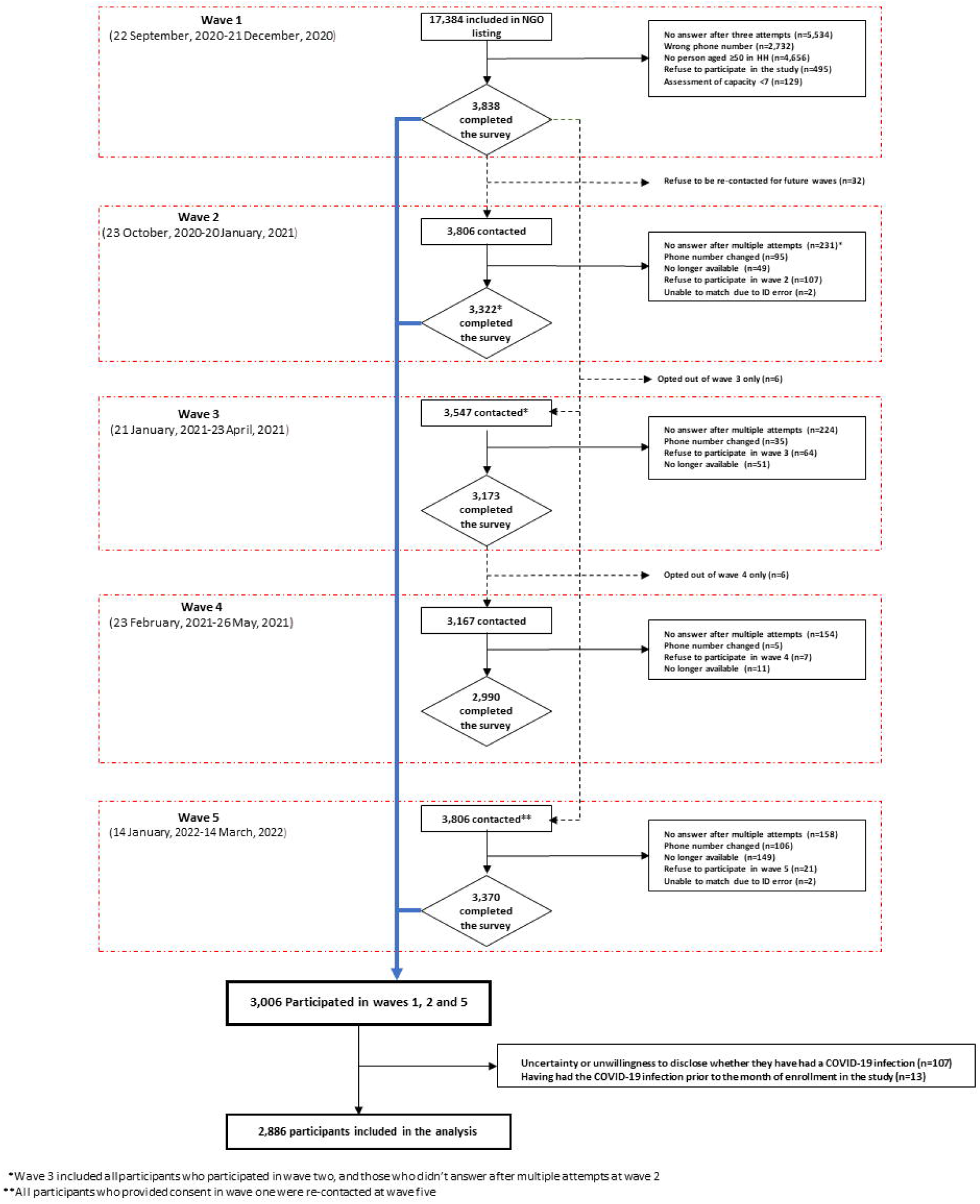
The flowchart of the study population

### 2.3. Data sources

The survey tool for each wave was created based on various sources, including established scales and context-specific questions and resulted from a collaborative effort engaging academics, humanitarian workers, government representatives, and refugee community members. The questionnaire was drafted in English, then translated into Arabic. Before collecting data, the Arabic version was tested internally with data collectors and local community focal points to ensure comprehension. The survey was conducted in Arabic via computer-assisted telephone interview by trained data collectors, and responses were entered using Kobotoolbox. Daily monitoring was conducted simultaneously with data collection to ensure the quality of the data. Different sets of questions or modules were included at different waves. Additional information about the development of the survey tool can be found in previously published papers.^20,21^ For the current analysis, demographics, health and self-reported COVID-19 infection characteristics were extracted respectively from waves one (September 22 - December 21, 2020); two (October 23, 2020 – January 20, 2021) and five (January 14-March 14, 2022).

### 2.4. Outcome measures

Self-reported COVID-19 infection was the outcome of interest. The following question was asked in wave 5: “Have you ever had COVID-19?”. In addition, the participants were asked about whether their diagnosis was confirmed through a PCR test or lateral flow test, the reasons for not undergoing testing, and other questions related to their most recent infection, which are detailed in the Supplementary Material table A.1.

### 2.5. Candidate predictors

The literature review guided the inclusion of the following candidate predictors for COVID-19 infection: age, sex, education, residence outside or inside informal tented settlements, marital status, number of self-reported physician diagnosed chronic conditions, household food insecurity (measured using the 8-item Food Insecurity Experience Scale; individuals are categorized as not food insecure if their score ranges between 0 and 3, and as food insecure if their score is 4 or higher),^22^ household water insecurity (measured using the short-form Household Water Insecurity Scale),^23^ having unmet waste management needs, receipt of cash assistance and having legal status documentation. The questions used to assess these predictors along with the corresponding survey wave from which they were obtained are provided in the Supplementary Material table A.2.

### 2.6. Missing data

The missing values did not exceed 5% for all predictors and were considered to be missing at random. Therefore, a complete case analysis was performed.

### 2.7. Statistical analysis

Descriptive analysis was carried out to report the frequencies and percentages for categorical variables and median and interquartile range for continuous variables. Unadjusted odds ratios

(OR) and 95% confidence intervals (95%CI) for each potential predictor were assessed and reported by using unadjusted logistic regressions. A two-sided p-value below 0.05 was considered as statistically significant.

All candidate predictors were categorical except age, which had a linear relationship with the outcome. The presence of multicollinearity was evaluated and a variance inflation factor above five indicated high collinearity. All candidate predictors of self-reported COVID-19 infection were entered into adaptive least absolute shrinkage and selection operator (Lasso) model. The lasso regression methods for predictor selection, applies a shrinkage effect on the coefficient estimates towards zero by selecting an optimal value for Lambda, using 10-fold cross-validation.^24^ The penalized coefficients were reported and used to calculate the predicted risk. Double-selection lasso logistic regression, which is a lasso for inference model,^25^ was used to provide ORs accompanied by their corresponding 95% CI.

The discriminatory ability of the models was assessed using the Area Under the Curve (AUC), which ranges from 0.5 to 1.0. A value of 1.0 indicates a perfect ability to distinguish between those with and without the outcome, while a value of 0.5 suggests a discriminatory ability equivalent to chance. The calibration of models, which measures the agreement between observed outcomes and the model’s predictions, was also evaluated, through the C-slope and calibration plot. The calibration plot categorizes individuals into 10 groups based on predictive probabilities and plots the mean predicted risk against the mean observed proportion of events. In a perfectly calibrated model, the plot would show a diagonal line with an intercept of 0 and a slope of 1. A C-slope less than 1 indicated that the model has been overfitted. Similarly, the quality of the prediction model is reflected by Brier score, ranging from 0 (indicating a perfect prediction accuracy) to 1 (indication the worst prediction accuracy). Furthermore, calibration-in-the-large (CITL) was evaluated to determine the overall disparity between the observed frequency of events and the average predicted risk. A sensitivity analysis was conducted by restricting participants to those with confirmed COVID-19 infection via PCR testing. Overall, the model that exhibited a calibration slope closest to one and achieved a higher AUC was selected as the optimal model. All analyses were conducted using Stata/SE 16.

### 2.8. Ethical approval

This study was approved by the American University of Beirut Social and Behavioral Sciences Institutional Review Board [Reference: SBS-2020-0329]. Verbal informed consent for participation was obtained from all participants.

## 3. Results

Out of the initial 17,384 households contacted during the first wave, 3,838 answered and were eligible, and a total of 3,322 Syrian refugees successfully completed both waves one and two. Among them, 3,006 participants also completed wave five. After excluding 120 participants who were uncertain or unwilling to disclose their history of COVID-19 infection or had experienced a COVID-19 infection prior to wave one, the final sample for analysis comprised 2,886 participants (Figure 1). The median age of the participants was 56 (IQR=52-62; range=50-96); 1,527 (52.9%) were males and 1,761 (61.0%) lived outside tented settlements (Table 1). Among the 2,886 participants, 283 individuals (9.8% [95%CI: 8.7% to 10.9%]) reported experiencing the COVID-19 infection at least once.

**Table 1:**
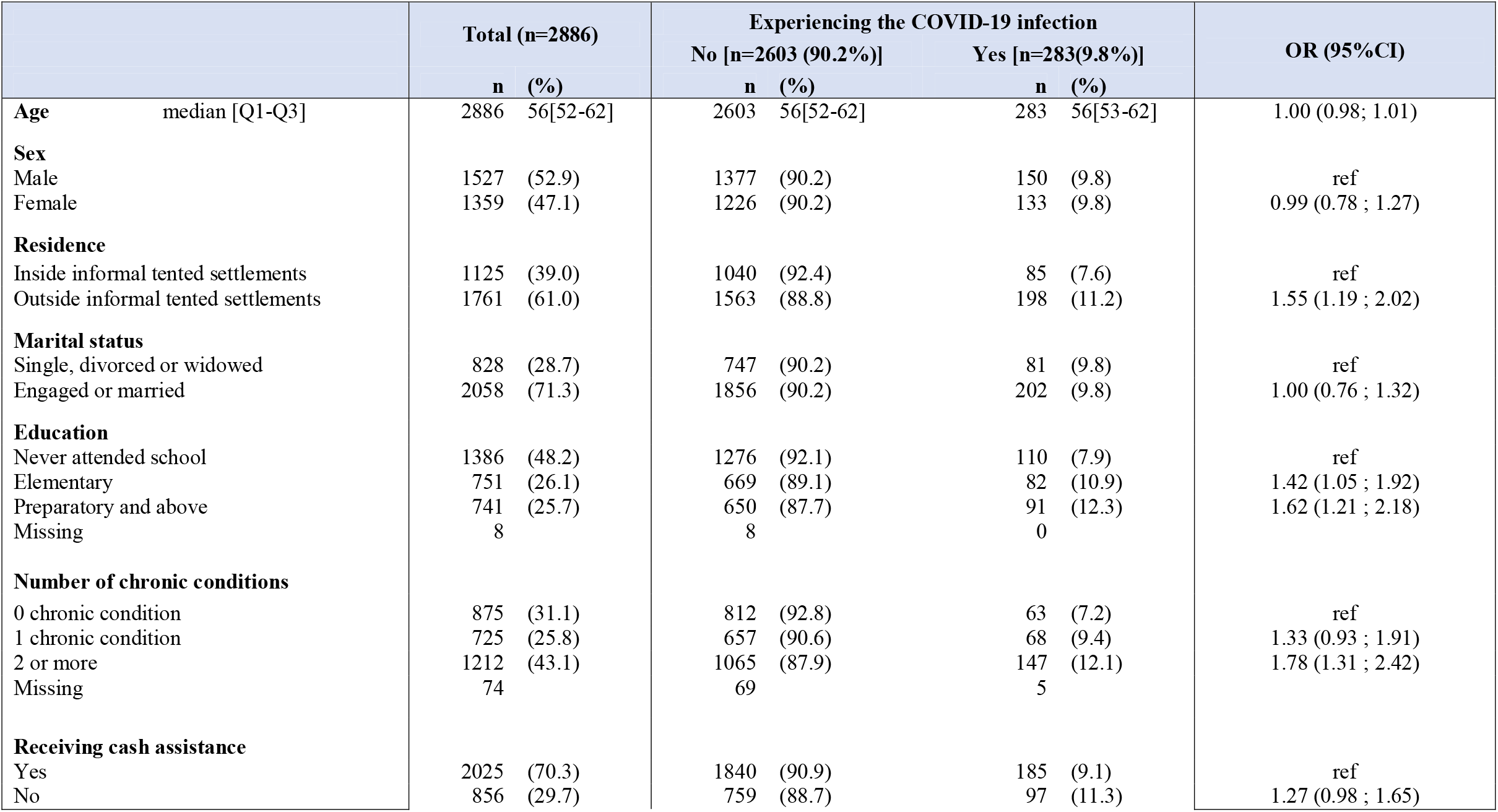
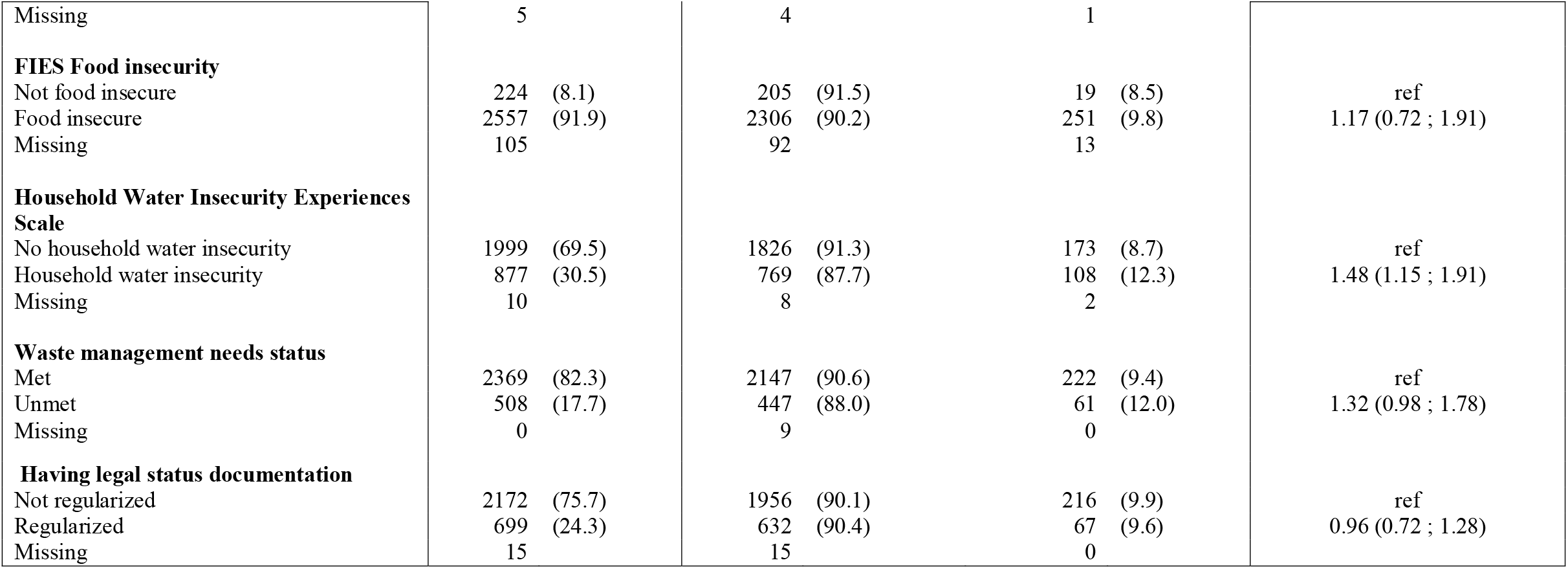
Characteristics of the study population and their association with the COVID-19 infection.

The bivariate analysis showed that living outside informal tented settlements (OR=1.55[95%CI: 1.19 to 2.02]) compared to living inside, having elementary (OR=1.42[95%CI: 1.05 to 1.92]) and preparatory education or above (OR=1.62[95%CI: 1.21 to 2.18]) compared to never having attended school, having at least two chronic conditions (OR=1.78[95%CI: 1.31 to 2.42]) compared to not having any, and experiencing household water insecurity (OR=1.48[95%CI: 1.15 to 1.91]) compared to household water security were associated with increased odds of COVID-19 infection. No statistically significant associations were found between the COVID-19 infection and age, sex, marital status, receiving cash assistance, household food insecurity, unmet waste management needs and legal status documentation (Table 1).

Six predictors of COVID-19 infection were identified including residency, education, number of chronic conditions, receiving cash assistance, household water insecurity and unmet waste management needs (Table 2). The model had a C-statistic of 0.621 (95%CI: 0.587 to 0.655), a calibration slope of 1.004 (95%CI:0.704 to 1.304), a brier score of 0.088 and CITL of 0.005 (95%CI: −0.121 to 0.131), which indicate moderate discriminative ability and good calibration. Figure 2 shows the calibration plot of the model and Table 2 presents the penalized coefficients, adjusted ORs, and their associated 95%CI.

**Table 2:** Predictive model of at COVID-19 infection.

| Predictors | Lasso for prediction* |  | Lasso for inference** |  |  |
| --- | --- | --- | --- | --- | --- |
|  | Codes | Coefficients | OR | (95% CI) | P-value |
| **Residency** |  |  |  |  |  |
| Inside informal tented settlements | 0 |  |  | *ref* |  |
| Outside informal tented settlements | 1 | 0.37 | 1.44 | (1.08 ; 1.91) | 0.012 |
| **Education** |  |  |  |  |  |
| Never attended school | 0 |  |  | *ref* |  |
| Elementary | 1 | 0.23 | 1.45 | (1.02 ; 2.06) | 0.036 |
| Preparatory and above | 2 |  | 1.64 | (1.15 ; 2.35) | 0.006 |
| **Number of chronic conditions** |  |  |  |  |  |
| 0 chronic condition | 0 |  |  | *ref* |  |
| 1 chronic condition | 1 | 0.27 | 1.34 | (0.92 ; 1.95) | 0.122 |
| 2 or more | 2 |  | 1.74 | (1.24 ; 2.44) | 0.001 |
| **Receiving cash assistance** |  |  |  |  |  |
| Yes | 0 |  |  | *ref* |  |
| No | 1 | 0.17 | 1.21 | (0.92 ; 1.59) | 0.161 |
| **Household Water Insecurity Experiences Scale** |  |  |  |  |  |
| No household water insecurity | 0 |  |  | *ref* |  |
| Household water insecurity | 1 | 0.37 | 1.45 | (1.11 ; 1.90) | 0.006 |
| **Waste management needs status** |  |  |  |  |  |
| Met | 0 |  |  | *ref* |  |
| Unmet | 1 | 0.23 | 1.29 | (0.94 ; 1.78) | 0.109 |
| **Constant** |  | -3.20 |  |  |  |

\*Lasso for prediction performed to report the coefficients used to calculate the predicted risk
\*\*Lasso for inference was used to report the odds ratio and 95% confidence interval

**Figure 2.**
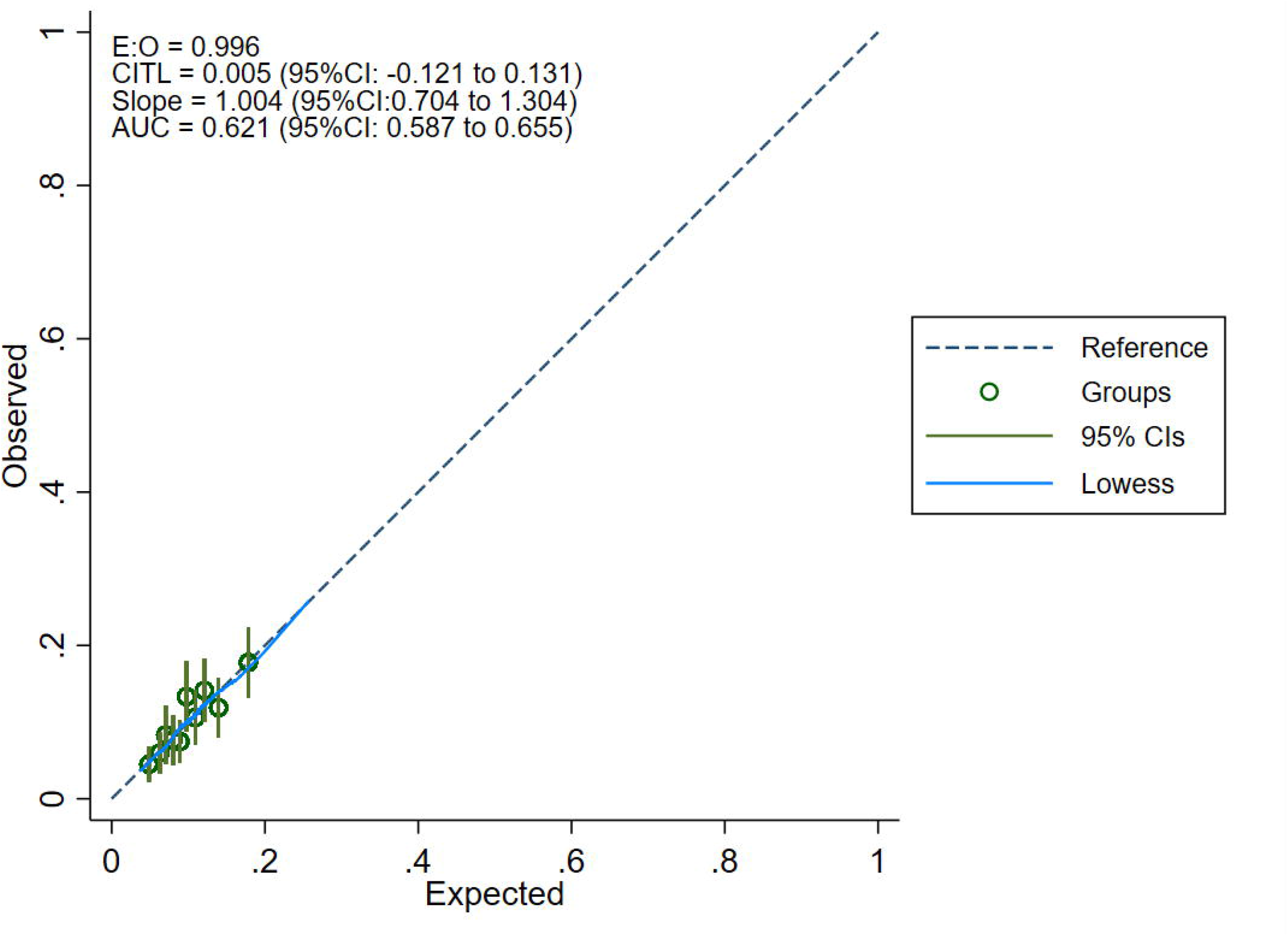
Calibration plot of the final model

In a sensitivity analysis, the prediction model of COVID-19 confirmed by PCR included residency, education level, and the number of chronic conditions (Supplementary Material table A.3). Although the model’s performance was acceptable, it is not better than the performance of the original model (C-Statistic:0.628 [0.581 to 0.674] and C-Slope:0.992 [0.587 to 1.397]) (Supplementary Material table A.3).

The predictors’ coefficients indicate that living outside informal tented settlements compared to living inside, having higher education, multimorbidity, not receiving cash assistance, household water insecurity and having unmet waste management needs increased the risk of COVID-19 infection.

To illustrate, the predicted risk could be calculated using the formula below (Table 2):

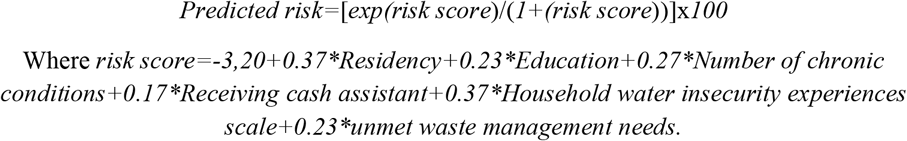

The predicted risk of having COVID-19 for a Syrian refugee who was living inside informal tented settlements, never attended school, did not have any chronic conditions, received cash assistance, was water secure and had waste management needs met was 3.9%. However, for another individual who was residing outside informal tended settlements, had preparatory education, two chronic conditions, did not receive cash assistance, was water insecure and had unmet waste management needs, was 25.7%.

Nearly half of participants who had COVID-19 infection (n=138[49.1%]) were diagnosed through PCR or lateral flow tests. The main reasons for not taking the tests were that they did not consider it necessary due to having COVID-19 symptoms (n=91[63.6%]), or they were unable to afford its cost (n=46[32.2%]) (Figure 3). No statistically significant associations were found between PCR testing and the participants’ characteristics (Supplementary Material table A.4). In addition, 21 participants (7.4%) had the infection more than twice from the start of the pandemic until March 2022 (Supplementary Material table A.5). The most recent infection among participants occurred during the period of November 2020 to March 2022, with peaks during February 2021 (n=30[10.6%]), December 2021 (n=32[11.3%]), and January 2022 (n=32[11.3%]) (Supplementary Material figure A.1).

**Figure 3.**
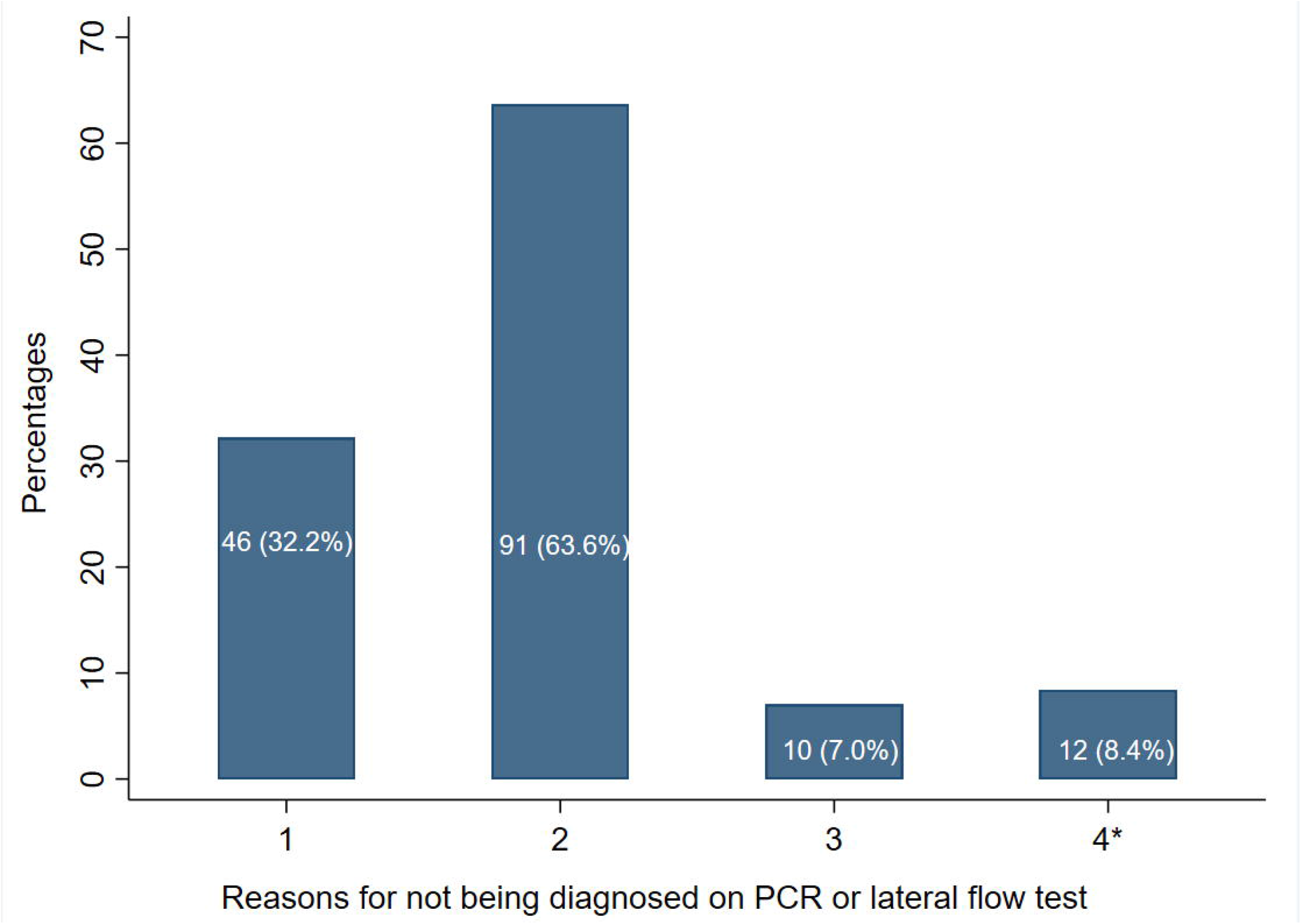
The reasons for not being diagnosed on PCR or lateral test.

During their last infection, 95.8% of participants stayed at home (n=271; median [IQR]: 3 weeks [2;4]), 90.8% wore a mask (n=257), 79.4% used traditional remedies (garlic, ginger, anise tea) (n=224), 73.1% took medication prescribed by the doctor (n=207), 53% called (n=152) or visited a doctor or healthcare centre (n=151) and 43.8% went to the pharmacy (n=124) (Supplementary Material table A.5).

## 4. Discussion

During the period of November 2020 and March 2022, the prevalence of COVID-19 infection was 9.8% in this population of older Syrian refugees (95%CI: 8.7% to 10.9%). The predictive model identified from this study had good calibration and moderate discriminative ability and included six predictors of the infection: residency, education, number of chronic conditions, receiving cash assistance, household water insecurity and unmet waste management needs. Nearly half of the cases were diagnosed through PCR or lateral flow tests. The primary reasons for not taking the test were the perception that it was unnecessary due to having symptoms or the inability to afford it. Overall, there was high adherence to preventive measures during the period of infection, in addition to the consumption of prescribed medication or traditional remedies.

This study showed that the prevalence of COVID-19 infection among older Syrian refugees in Lebanon was 9.8%. A small-sample telephone survey which was similarly conducted in Lebanon by the Access Center for Human Rights, found that one-third of the 217 Syrian refugees contacted in April 2021 reported experiencing COVID-19 symptoms, most of whom were less than 44 years old. In addition, two nationwide population-based cross-sectional studies reported the prevalence of COVID-19 infection in Lebanon. The first study was based on a self-reported outcome and found a prevalence of 13.9% out of 1,119 participants, with a higher risk of infection among older adults.^26^ The second study was based on lateral flow test and found a crude seroprevalence of 16.0%, with a slightly higher prevalence among those aged 60 years or older (18.5%). However, no statistically significant difference was found between the age groups.^27^ The timeline of the most recent COVID-19 infection in this study is almost aligned with the epidemic curve of confirmed cases.^2^ These data showed that the prevalence of COVID-19 infection from our study of older Syrian refugees was lower than the national prevalence and could be due to an underestimation resulting from the lack of testing among this population. This pattern of underrepresentation of the true disease burden among displaced populations during pandemics was also observed in previous outbreaks beyond COVID-19.^28^

Globally, displaced populations frequently encounter individual, cultural, environmental, and financial barriers to access healthcare services,^29,30^ including testing for infectious diseases during pandemics.^31,32^ Particularly, Syrian refugees in Lebanon faced barriers to PCR testing which were mainly related to individuals’ perceptions of lack of need or lack of affordability,^14,26^ despite the availability of free testing services and the national effort to raise awareness.^14^ Perceived stigma, discrimination, fear of deportation, and limited accessibility to testing centers were reported as additional barriers to testing among Syrian refugees in Lebanon,^14^ and other displaced populations.^31–35^ Given that nearly half of the participants visited doctors, healthcare services or pharmacies after getting infected, the implementation of COVID-19 screening in primary healthcare centers and pharmacies, could be an important strategy to control the spread of infection among Syrian refugees, as these settings are trusted sources and the frontlJline of healthcare services.^14^ In addition, in future pandemics, due to the use of non-prescribed medications and traditional remedies reported by older Syrian refugees, it will be important for humanitarian organizations to intensify their health promotion programs to increase awareness of testing availability and the importance of visiting a healthcare professional.

This study, focusing on older Syrian refugees in Lebanon, identified six predictors of COVID-19 infection with acceptable predictive abilities. These factors primarily pertain to socio-economic and health aspects, previously recognized in predictive models, such as education^15^ and the number of chronic conditions.^15,16^ Residency and receiving cash assistance emerged as unique factors within the refugee context. Notably, there is no prior research incorporating unmet waste management needs or water insecurity into prediction models, highlighting the added value of this study to the literature.

Contrary to previous findings higher level of education has been regarded as a protective factor against COVID-19,^36^ while in this study higher education had an increased odds of COVID-19 infection. During the pandemic, most Syrian refugees lost their jobs either permanently or temporarily, particularly those working in semi-skilled and unskilled labour such as agriculture and construction.^37,38^ In contrast, individuals in more skilled occupations, which are typically associated with higher education,^37^ were less likely to be laid-off. Consequently, Syrian refugees with higher education were likely to have had jobs that required social interactions, which increased their risk of COVID-19 infection (Supplementary Material table A.6).

Having chronic conditions was identified as another predictor of COVID-19 infection. A systematic review examining the risk factors of COVID-19 in migrant populations suggested the presence of comorbidities could be associated with a higher risk of infection.^39^ This connection could be attributed to that individuals with chronic conditions often need to visit healthcare centers and hospitals for management, therefore increasing their risk of exposure to the virus.^39^

Insecurities in basic needs, such as water insecurity and unmet waste management needs, could contribute to the spread of viruses including COVID-19 infection, through various pathways.^15,40^ These include inadequate access to safe drinking water, drinking insufficient quantity of water, and limitations in practicing essential hygiene measures such as handwashing, bathing, and cleaning surfaces.^41^ It is crucial to emphasize that the majority of Syrian refugees rely on humanitarian assistance to meet their essential needs, including water and sanitation.^12^ Consequently, the absence of this assistance could worsen their living conditions, and create opportunities for viral transmission within their households and communities.

Syrian refugees residing in informal tented settlements may be more easily reached by NGOs compared to those living in host communities.^42^ As a result, reach of humanitarian programs including cash assistance, awareness campaigns, masks, hygiene kits and mobile clinics could reduce their risk of contracting the infection. These findings reinforce our previous recommendation, emphasizing the need for humanitarian organizations to redesign and expand their programs to ensure coverage for all refugees, including those living outside informal tented settlements, in order to address inequities and effectively control the pandemic.^20^

Given the limited number and social aspect of determinants identified for predicting infection, the model could serve as a quick, effective, and cost-efficient tool for humanitarian organizations to identify refugees at a higher risk of contracting the virus in future pandemics and outbreaks. Researchers and agencies are strongly encouraged to assess the model’s performance across diverse humanitarian contexts. In addition, existing secondary data from previous major infectious disease outbreaks affecting displaced populations, such as cholera, measles could be used to validate the model, to ascertain its generalizability beyond COVID-19 and its predictive power across different epidemiological scenarios and contexts.

This study has several limitations, firstly, the self-reported data could lead to a risk of social desirability bias, whereby refugees could be overestimating adherence to preventive measures. Additionally, COVID-19 infection could be underreported due to their fear of stigma, leading to possible misclassification bias. Additional predictors for COVID-19 infection could be added to the model in order to improve its performance, but they were not included due to their time-dependent and context-specific nature. Furthermore, the study sample represents beneficiaries from a specific humanitarian organization and may not be representative of all Syrian refugees in Lebanon.

## 5. Conclusions

In conclusion, the identified predictors could be used to target Syrian refugees at higher risk of COVID-19 infection and future infectious disease outbreaks particularly those who require to work, seek healthcare or live outside tented settlements. The provision of humanitarian assistance is crucial to improve living conditions and reduce vulnerability to infectious diseases in future pandemics. Prediction models that incorporate social determinants offer a practical tool for humanitarian and public health actors to identify at-risk individuals early and allocate resources more efficiently. Integrating such models into emergency preparedness plans can strengthen anticipatory action and enable rapid targeting of vulnerable populations. Furthermore, intensifying awareness campaigns and considering the implementation of screening measures in the community are essential to control the spread of the virus in this population.

## Data Availability

The deidentified data could be shared upon request from NRC and AUB.

## 6. List of abbreviations

UNHCR: UN High Commissioner for Refugees
MENA: Middle East and North Africa
PCR: Polymerase Chain Reaction
NGO: Non-Governmental Organization
OR: Odds Ratios
95%CI: 95% confidence intervals
LASSO: least absolute shrinkage and selection operator
AUC: Area under the curve
CITL: Calibration-in-the-large

## 7. Declarations

### Ethical approval

This study was approved by the American University of Beirut Social and Behavioral Sciences Institutional Review Board [Reference: SBS-2020-0329]. Verbal consent for participation was obtained from all participants.

### Consent for publication

Not applicable

### Availability of data and materials

The deidentified data could be shared upon request from NRC and AUB.

### Authors contributions

SA, HG, and SM conceptualized the study, obtained the primary funding for the study and designed the survey. SM obtained supplemental funding for this specific manuscript. SM, HG, SA, BA and TK contributed to data collection. BA contributed to the literature search and drafted the manuscript. The following drafts were reviewed and revised by SM, HG, BA and TK. The underlying survey data were verified by SM and BA. SM supervised BA and TK throughout the project. All authors have reviewed and approved the final version of the paper.

## Acknowledgments

The authors would like to thank Mrs. Noura Salibi and Miss Maria El Haddad for their support and expertise throughout all aspects of the research study, including data monitoring, cleaning and analysis. The authors would also like to acknowledge B.O.T (Bridge. Outsource. Transform) for their assistance in collecting the data required for the success of this study and the study participants for their participation.

## 8. Captions for figures

Labels: (1) Unable to afford PCR test/lateral flow; (2) I didn’t think the test was necessary because I had covid-19 symptoms; (3) I didn’t think the test was necessary because I had close contact with someone who had COVID-19; (4) Others; *Others include inability to access PCR testing centre, fear of diagnosis and absence of recommendation from doctors or healthcare professionals.

## 9. Online Supplementary Material Content

Table A.1: The survey questions on COVID-19 infection

Table A.2: The survey questions for the candidate predictors

Table A.3: Prediction model of COVID-19 infection, including participants diagnosed through PCR (n=2741)

Table A.4: Association between the characteristics of participants who had a COVID-19 infection and PCR testing

Table A.5: Description of participant behaviours following the COVID-19 infection Figure A.1: Last occurrence of COVID-19 infection

