## Supplementary material for "Predicting COVID-19 Infection Among Older Syrian Refugees in Lebanon to inform outbreak preparedness: A Multi-Wave Survey"

### **Table A.1: The survey questions on COVID-19 infection**

| **Have you ever had COVID-19?** | |
| --- | --- |
| - Yes - No - Don’t know - Refuse to answer | |
| **If yes, the following questions were asked** | |
| How many times have you had COVID-19? |  |
| When was the last time you have COVID-19? | Month / Year |
| Think of the last time you had COVID-19, when you found out, which of the measures did you take? *(please select all that apply)* ? | - Stay at home - Call doctor - Visit healthcare center or doctor - Go to the pharmacist - Wear a mask - Take medication prescribed by the doctor - Take medication on your own without prescription - Call UNHCR - Call MOPH hotline - Use traditional remedies (garlic, ginger, yensoon) - Other |
| - If other, specify | ……………………………. |
| - If stay at home – how long did you stay after you found out you had COVID-19? | Length of time:  Days…………….  Weeks………….. |
| If ever diagnosed with covid-19, was this diagnosed using either a PCR or lateral flow test? | - Yes - No - Don’t know / Refuse to answer |
| - If no, select the reason(s) why | - Unable to afford PCR test/lateral flow - Unable to access PCR testing center - I didn’t think the test was necessary because I had covid-19 symptoms - I didn’t think the test was necessary because I had close contact with someone who had COVID-19 - Fear of diagnosis - Other, Specify ………………………….. |

### **Table A.2: The survey questions for the candidate predictors**

| **Possible predictors** | **Wave*** | **Questions** |
| --- | --- | --- |
| Age | Wave 1 | Confirm participant’s year of birth |
| Sex | Wave 1 | Confirm participant’s gender |
| Residency | Wave 1 | Confirms residence inside/outside ITS |
| Education | Wave 1 | Have you ever attended school?  If yes, what is your level of education? [elementary, preparatory, secondary, vocational, university and post graduate] |
| Marital status | Wave 1 | What is your marital status? [single, engaged, married, separated/divorced, widowed] |
| Number of chronic disease conditions | Wave 2 | Have you ever been told by a health care professional that you have any of the following chronic illnesses? (select all that apply): Hypertension, Diabetes, Cardiovascular Diseases, Chronic Respiratory Diseases, Rheumatoid Arthritis, Chronic kidney diseases and Cancer |
| Food insecurity | Wave 1 | Food Insecurity Experience Scale^1^ |
| Water insecurity | Wave 1 | Household Water Insecurity Experiences Scale^2^ |
| Unmet waste need | Wave 2 | Do you have an unmet waste need? |
| Receipt of cash assistance | Wave 1 | Have you received any cash assistance in the last 6 months? |
| Regularization | Wave 2 | Do you have regularized residency in Lebanon? |

* Wave 1: September-December 2020; Wave 2: October 2020 – January 2021

1 Cafiero C, Viviani S, Nord M. Food security measurement in a global context: The food insecurity experience scale. *Measurement.* 2018;116:146-152.

2 Young SL, Miller JD, Frongillo EA, Boateng GO, Jamaluddine Z, Neilands TB. Validity of a four-item household water insecurity experiences scale for assessing water issues related to health and well-being. *The American journal of tropical medicine and hygiene.* 2021;104(1):391.

### **Table A.3: Prediction model of COVID-19 infection, including participants diagnosed through PCR (n=2741)**

| **Predictors** | **Lasso for prediction*** | | **Lasso for inference**** | | |
| --- | --- | --- | --- | --- | --- |
|  | **Codes** | **Coefficients** | **OR** | **(95% CI)** | **P-value** |
| **Residency** |  |  |  |  |  |
| Inside informal tented settlements | 0 | 0.46 | *ref* | | 0.035 |
| Outside informal tented settlements | 1 |  | 1.53 | (1.03 ; 2.28) |  |
| **Education** |  |  |  |  |  |
| Never attended school | 0 | 0.15 | *ref* | |  |
| Elementary | 1 |  | 1.67 | (1.04 ; 2.68) | 0.034 |
| Preparatory and above | 2 |  | 1.62 | (0.97 ; 2.69) | 0.064 |
| **Number of chronic conditions** |  |  |  |  |  |
| 0 chronic condition | 0 | 0.43 | *ref* | |  |
| 1 chronic condition | 1 |  | 0.99 | (0.55 ; 1.77) | 0.980 |
| 2 or more | 2 |  | 2.01 | (1.24 ; 3.25) | 0.004 |
| **Constant** |  | -3.89 |  |  |  |
| **Model performance** | **Estimates** | **[95% CI]** |  |  |  |
| C-statistic | 0.628  (0.581; 0.674) | |  |  |  |
| C-slope | 0.992  (0.587; 1.397) | |  |  |  |
| CITL | 0.017 (-0.155; 0.190) | |  |  |  |

*Lasso for prediction performed to report the coefficients used to calculate the predicted risk

**Lasso for inference was used to report the odds ratio and 95% confidence interval

### **Table A.4: Association between the characteristics of participants who had a COVID-19 infection and PCR testing**

|  | **Diagnosed on PCR or lateral flow test** | | | | **OR (95%CI)** |
| --- | --- | --- | --- | --- | --- |
|  | **No (n=143)** | | **Yes (n=138)** | |  |
|  | **n** | **(%)** | **n** | **(%)** |  |
| **Age Median [IQR]** |  | 55 [52 ;61] |  | 58 [53 ;63] | 1.03 (0.99 ; 1.06) |
| **Gender** |  |  |  |  |  |
| Male | 83 | (55.7) | 66 | (44.3) | ref |
| Female | 60 | (45.5) | 72 | (54.5) | 1.51 (0.94 ; 2.42) |
| **Residence** |  |  |  |  |  |
| Inside informal tented settlements | 46 | (54.1) | 39 | (45.9) | ref |
| Outside informal tented settlements | 97 | (49.5) | 99 | (50.5) | 0.83 (0.50 ; 1.38) |
| **Marital status** |  |  |  |  |  |
| Single, divorced or widowed | 38 | (47.5) | 42 | (52.5) | ref |
| Engaged or married | 105 | (52.2) | 96 | (47.8) | 0.83 (0.49 ; 1.39) |
| **Education** |  |  |  |  |  |
| Never attended school | 52 | (47.7) | 57 | (52.3) | ref |
| Elementary | 40 | (48.8) | 42 | (51.2) | 0.96 (0.54 ; 1.70) |
| Preparatory and above | 51 | (56.7) | 39 | (43.3) | 0.70 (0.40 ; 1.22) |
| **Number of chronic conditions** |  |  |  |  |  |
| 0 chronic condition | 33 | (53.2) | 29 | (46.8) | ref |
| 1 chronic condition | 42 | (61.8) | 26 | (38.2) | 0.70 (0.35 ; 1.42) |
| 2 or more | 64 | (43.8) | 82 | (56.2) | 1.46 (0.80 ; 2.65) |
| **Receiving cash assistance** |  |  |  |  |  |
| Yes | 88 | (48.1) | 95 | (51.9) | ref |
| No | 54 | (55.7) | 43 | (44.3) | 0.74 (0.45 ; 1.21) |
| **FIES Food insecurity** |  |  |  |  |  |
| Not food insecure | 7 | (36.8) | 12 | (63.2) | ref |
| Food insecure | 131 | (52.6) | 118 | (47.4) | 0.52 (0.20 ; 1.38) |
| **Household Water Insecurity Experiences Scale** |  |  |  |  |  |
| No household water insecurity | 81 | (47.4) | 90 | (52.6) | ref |
| Household water insecurity | 61 | (56.5) | 47 | (43.5) | 0.69 (0.43 ; 1.12) |
| **Unmet waste need** |  |  |  |  |  |
| No | 113 | (51.1) | 108 | (48.9) | ref |
| Yes | 30 | (50.0) | 30 | (50.0) | 1.05 (0.59 ; 1.85) |
| **Regularization** |  |  |  |  |  |
| No | 107 | (50.0) | 107 | (50.0) | ref |
| Yes | 36 | (53.7) | 31 | (46.3) | 0.86 (0.50 ; 1.49) |

### **Table A.5: Description of participant behaviours following the COVID-19 infection**

|  | **n** | **(%)** |
| --- | --- | --- |
| **How many times have you had COVID-19?** |  |  |
| 1 | 262 | (92.6) |
| 2 | 17 | (6.0) |
| 3 | 4 | (1.4) |
| **Diagnosed on PCR or lateral flow test** |  |  |
| No | 143 | (50.9) |
| Yes | 138 | (49.1) |
| **Measures taken the last time you had COVID-19** |  |  |
| Stay at home | 271 | (95.8) |
| Wear a mask | 257 | (90.8) |
| Use traditional remedies (garlic, ginger, anise) | 224 | (79.4) |
| Take medication prescribed by the doctor | 207 | (73.1) |
| Call doctor | 152 | (53.7) |
| Visit healthcare center or doctor | 151 | (53.4) |
| Go to the pharmacist | 124 | (43.8) |
| Take medication on your own without prescription | 61 | (21.6) |
| Call UNHCR | 46 | (16.3) |
| Call MOPH hotline | 16 | (5.7) |
| **How long did you stay at home after you found out you had COVID-19? (weeks)** Median [Q1-Q3] | 271 | 3 [2;4] |

**Table A.6: Educational level by work for pay during the last 7 days (wave 1)**

| **Education** | **Work for pay during the last 7 days** | | **P-value** |
| --- | --- | --- | --- |
|  | **No**  n (%) | **Yes**  n (%) |  |
| Never attended school | 1271 (91.7) | 115 (8.3) | P<0.001 |
| Elementary | 645 (85.9) | 106 (14.1) |  |
| Preparatory and above | 631 (85.2) | 110 (14.8) |  |

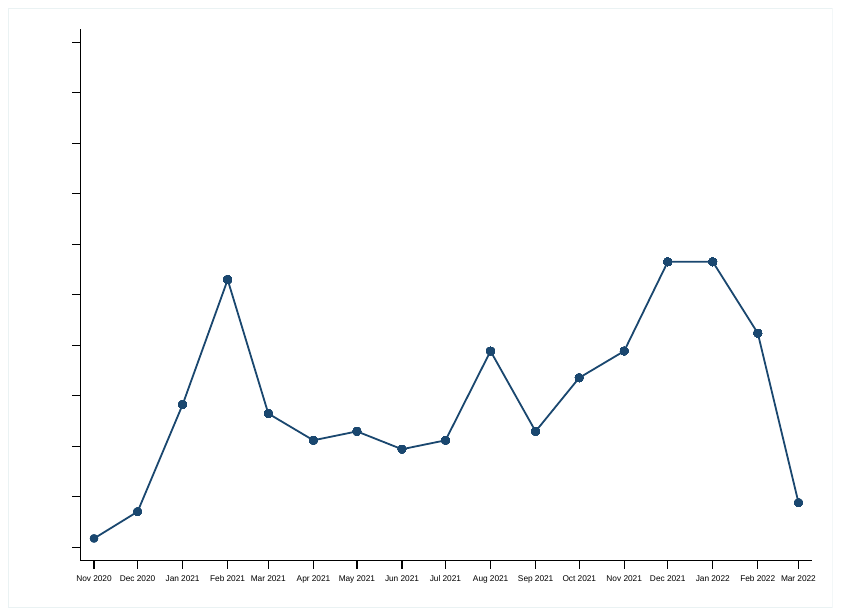

### **Figure A.1: Last occurrence of COVID-19 infection**
